# Bibliometric Analysis of Global Scientific Research on SARS-CoV-2 (COVID-19)

**DOI:** 10.1101/2020.03.19.20038752

**Authors:** Fatemeh Rafiei Nasab, Fakher Rahim

## Abstract

**Background and Aim:** Since late 2019, an unknown-origin pneumonia outbreak detected in Wuhan city, Hubei Province, China. We aimed to build a model to qualitatively and quantitatively assess publications of research of COVID-19 from 2019 to 2020.

**Materials and Methods:** Data were obtained from the Web of Science (WOS), PubMed, and Scopus Core Collection on March 02, 2020, and updated on March 10. We conducted a qualitative and quantitative analysis of publication outputs, journals, authors, institutions, countries, cited references, keywords, and terms according to bibliometric methods using VOS viewer c software packages.

**Results:** Initially, we identified 227 papers, of which after an exclusion process, 92 studies were selected for statistical analyses. China accounted for the highest proportion of published research (44 papers, 40.48%), followed by the United States (21 papers, 19.32%), and Canada (7 papers, 6.44%). Adjusted by gross domestic product (GDP), ranked first, with 0.003 articles per billion GDP. In total, the top 10 journals published 47 articles, which accounted for 51.08% of all publications in this Feld. A total of 6 studies (05.52%) were supported by National Natural Science Foundation of China. Chinese Academy of Sciences ranked second 2, 2.76%).

**Conclusion:** Bibliometric and visualized mapping may quantitatively monitor research performance in science and present predictions. The subject of this study was the fast growing publication on COVID-19. Most studies are published in journals with very high impact factors (IFs) and other journals are more interested in this type of research.

**Highlights:**

1. Bibliometric description and mapping provided a birds-eye view of information on Covid-19 related research
2. Readers to comprehend the history of published Covid-19 articles in just a few minutes.
3. We evaluated the research strength of countries and institutions,
4. Scholars might refer to in order to find cooperative institutions.
5. During our research using the selected database, we tried to guarantee comprehension and objectivity.

## Introduction

Since late 2019, an unknown-origin pneumonia outbreak detected in Wuhan city, Hubei Province, China [1]. Though the outbreak of corona virus disease 2019, SARS-CoV-2 (COVID-19) occurred in China first, it rapidly spread globally. Today, 146 countries and territories are reporting confirmed and suspected cases of COVID-19 [2]. Numerous etiological studies have conducted to find the detailed biological features of COVID-19 so far. Following the global outbreak and spread of the virus, the World Health Organization (WHO) issued a statement on January 11, 9191, announcing the outbreak of the COVID-19 as the sixth major public health emergency worldwide [3]. Thus, to prevent the spread of this new coronavirus, it is necessary health-care staffs and decision maker, governments and the people to work together. Moreover, it is recommended that all potentially exposed subjects to be isolated for 14 days, suggesting isolation is the best way to contain this epidemic [4].

Extracting information from basic and clinical research related to COVID-19 could be crucial for the improvement and development of the diagnosis, treatment and preventive strategies against this viral infection. A large number of epidemiological and clinical evidences have emerged, and a great deal of research has been published as well. Bibliometric studies is a method of investigating scientific achievements in a particular field of science through secondary analysis of the knowledge of published articles; so, it can help researchers appreciate the earlier and existing statuses of COVID-19 efficiently, predict and choose forthcoming advance directions, and design upcoming research [5, 6]. However, up to now, bibliometric studies on COVID-19 include inherent limitations and are rare. Due to the rapid growth of publishing articles in this field, the aim of this article is to assess the publication pattern of Covid-19 research globally. This study systematically assessed the publication distribution, stratified by geography, institution, funding sponsors, journals, and more. We also assessed the frequency of keywords and then employed Bibliometric mapping tools to demonstrate developments on Covid-19. Results were analyzed to further understand the structure of this field and to anticipate developments on Covid-19 research. Furthermore, this study can provide information for funding agencies to establish elated guidelines on Covid-19 research.

## Materials and Methods

### Sources of the Data and Search Strategy

The data in this article were based on the Web of Science (WOS), PubMed, and Scopus from 2019 to 2020. An initial comprehensive online search was performed on a single day, March 02, 2020, to avoid daily updating bias since the database is still open, and updated on March 10. The search key words were referred to MESH terms from PubMed and then were used as follows: TITLE-ABS-KEY(“Wuhan coronavirus”) OR TITLE-ABS-KEY(“Wuhan seafood market pneumonia virus”) OR TITLE-ABS-KEY(“COVID19 virus”) OR TITLE-ABS-KEY(“COVID-19 virus”) OR TITLE-ABS-KEY(“coronavirus disease 2019 virus”) OR TITLE-ABS-KEY(“SARS-CoV-2”) OR TITLE-ABS-KEY(“SARS2”) OR TITLE-ABS-KEY(“2019-nCoV”) OR TITLE-ABS-KEY(“2019 novel coronavirus”) OR TITLE-ABS-KEY(“2019 novel coronavirus infection”) OR TITLE-ABS-KEY(“COVID19”) OR TITLE-ABS-KEY(“coronavirus disease 2019”) OR TITLE-ABS-KEY(“coronavirus disease-19”) OR TITLE-ABS-KEY(“2019-nCoV disease”) OR TITLE-ABS-KEY(“2019 novel coronavirus disease”) OR TITLE-ABS-KEY(“2019-nCoV infection”) OR TITLE-ABS-KEY(“covid 19”) AND (LIMIT-TO (PUBYEAR, 2020) OR LIMIT-TO (PUBYEAR, 2019). Regarding manuscript types, only original articles and other document types were included. Ethical approval was not necessary, since the data were downloaded from the public databases and did not involve any interactions with human or animal subjects.

### Data Collection

The txt data download was imported into Microsoft Excel and VOS viewer. Bibliometric indicators were extracted from the data, including publication number, citation frequency, and H index. the data were analyzed both quantitatively and qualitatively. To adjust for economic condition and population size, statistics on gross domestic product (GDP) and population sizes from the Word Bank and the Central Intelligence Agency for the most recent report were used in the study. H-index is calculated as a measure of scientific research impact that reflects both the number of publications and the number of citations per publication: a scholar has published h papers, each of which has been cited in other papers at least h times.

### Statistical analysis

VOS viewer was used to analyze the relations among highly cited references and productive authors. It is commonly used for mapping and clustering of co citation network analysis. It also clusters citation terms and portrays the key words by color. The density of occurrence of information is portrayed by the size of the circle.(5)

## Results

### Countries Contributing to Global Publications

Initially, 227 papers were retrieved, dating back to 2019. After an exclusion process (**Figure 1**), 92 studies were selected for statistical analyses. **Figure 2** shows that articles in all countries and top three countries on Covid-19 researches.

**Figure 1:**
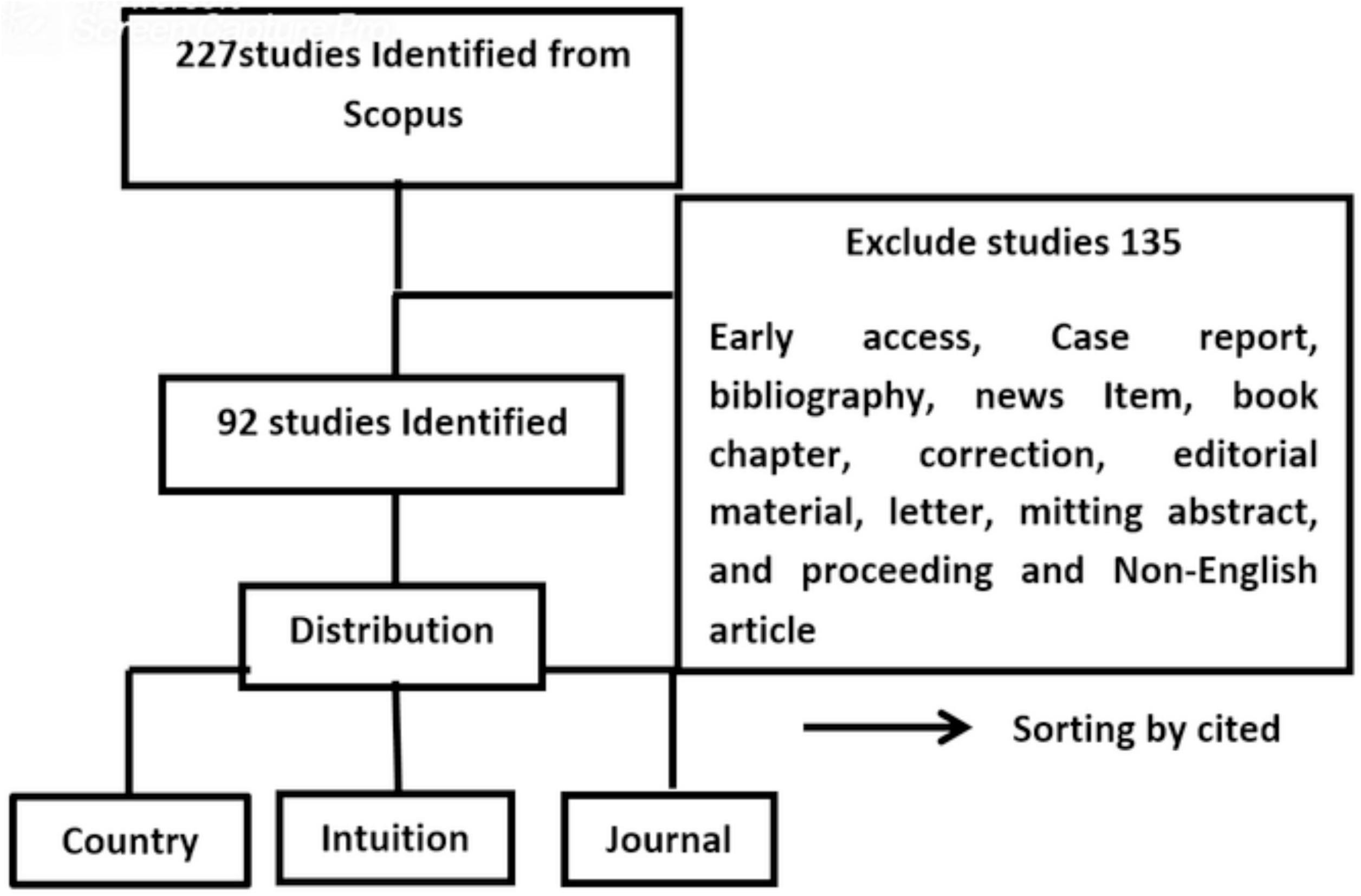
the inclusion and exclusion process of COVID-19 Research.

**Figure 2:**
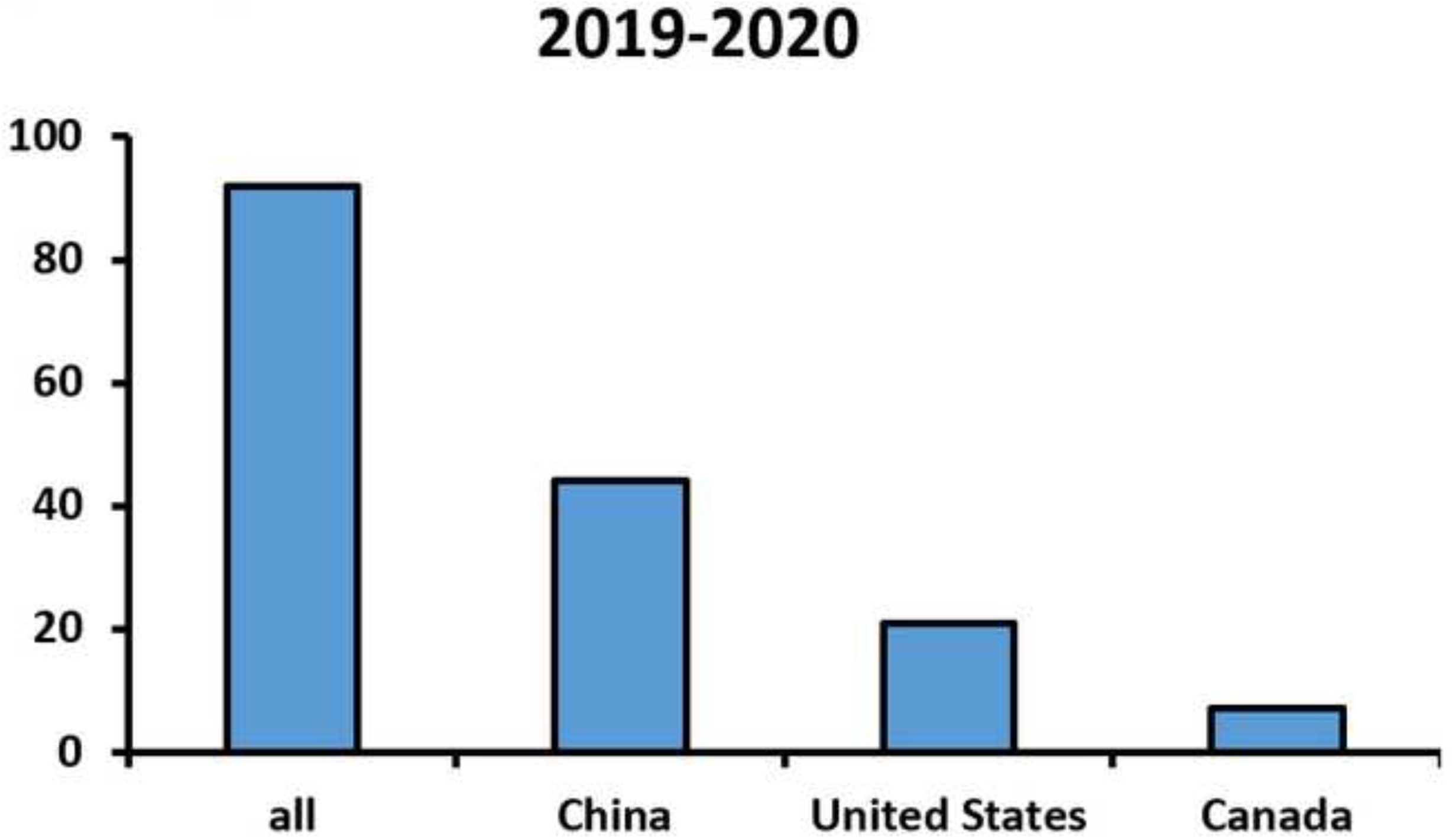
the number of worldwide and the top 3 countries publications on Covid-19 research

In terms of the most productive countries, China accounted for the highest proportion of published research (44 papers, 40.48%), followed by the United States (21 papers, 19.32%), and Canada (7 papers, 6.44%). Adjusted by gross domestic product (GDP), ranked first, with 0.003 articles per billion GDP. Adjusted by population, United States came to the fore with United States articles per million populations (**Table 1**).

**Table 1:** the 10 most productive countries related to COVID-19 research.

| country/territory |  | Percentage | N per million<br>population | N per billion<br>GDP | Total<br>citation | h-index |
| --- | --- | --- | --- | --- | --- | --- |
| China | 44 | 40.48 | 3.15 | 0.003 | 327 | 8 |
| United States | 21 | 19.32 | 6.41 | 0.001 | 19 | 2 |
| Canada | 7 | 6.44 | 1.88 | 0.004 | 1 | 1 |
| France | 6 | 5.52 | 8.95 | 0.002 | 9 | 1 |
| United<br>Kingdom | 6 | 5.52 | 9.02 | 0.002 | 8 | 1 |
| Italy | 5 | 4.6 | 8.27 | 0.016 | 5 | 1 |
| South Korea | 4 | 3.68 | 7.74 | 0.002 | 4 | 1 |
| Taiwan | 4 | 3.68 | 1.69 | 0.004 | 1 | 1 |
| Brazil | 3 | 2.76 | 1.43 | 0.001 | 5 | 1 |
| Netherlands | 3 | 2.76 | 1.74 | 0.003 | 10 | 1 |

Citation and H-Index Analysis. According to the analysis of the Scopus database, all articles related to Covid-19 had been cited a total of 364 times, an average of 3.95 times per paper. Specifically, the top 10 Covid-19 studies (with the highest citation frequency) accounted for 307 citations (84.34% of 364). In terms of countries, the China has the most citations (327) and the highest H-index (8). United States ranked second with 19 citations and an H-index of 2 (**Table 1**).

### Distribution of Institutes Focusing on Covid-19 research

The University of Hong Kong, Huazhong University of Science and Technology and Zhongnan Hospital of Wuhan University with six articles were top of the list. The top productive institutions can be viewed in the **Figure 3**.

**Figure 3:**
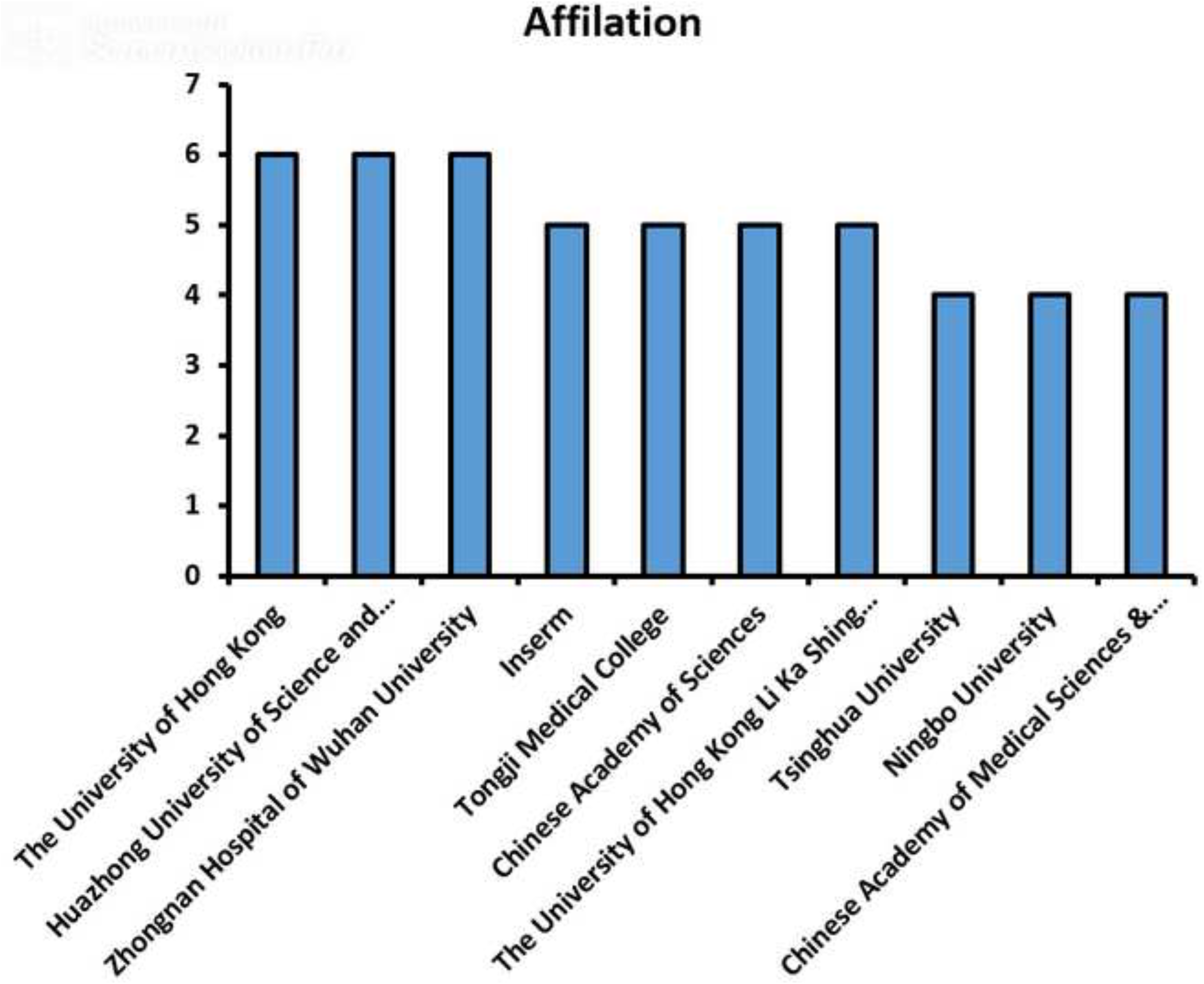
the number of publications on **COVID-19** research from the top 10 contribution institutes

### Distribution of Published Journals and Funding Agencies Focusing on Covid-19

Euro Surveillance has the greatest number of publications on Covid-19 Research with 10 papers. Journal of Medical Virology and Lancet Journal were on the second level either with 7 papers (**Figure 3**). In total, the top 10 journals published 47 articles, which accounted for 51.08% of all publications in this Feld. The top 10 funding bodies are shown in **Table 2**. A total of 6 studies (05.52%) were supported by National Natural Science Foundation of China. Chinese Academy of Sciences ranked second 2, 2.76%).

**Table 2:** Top 10 related funding sponsor

| Funding Sponsor | N | % |
| --- | --- | --- |
| National Natural Science Foundation of China | 6 | 5.52 |
| Chinese Academy of Sciences | 3 | 2.76 |
| National Basic Research Program of China (973 Program) | 3 | 2.76 |
| Chinese Academy of Medical Sciences | 2 | 1.84 |
| Fudan University | 2 | 1.84 |
| Ningbo University | 2 | 1.84 |
| Sanming Project of Medicine in Shenzhen | 2 | 1.84 |
| Science and Technology Major Project of Guangxi | 2 | 1.84 |
| Beijing Science and Technology Planning Project | 1 | 0.92 |
| Bill and Melinda Gates Foundation | 1 | 0.92 |

Most of the subject areas in the publication of the article on Covid-19 are as follows: Medicine (79 papers, 52.0%), Immunology and Microbiology (39 papers, 25.7%) and Biochemistry, Genetics and Molecular biology (9 papers, 5.9%) (**Figure 4**)

**Figure 4:**
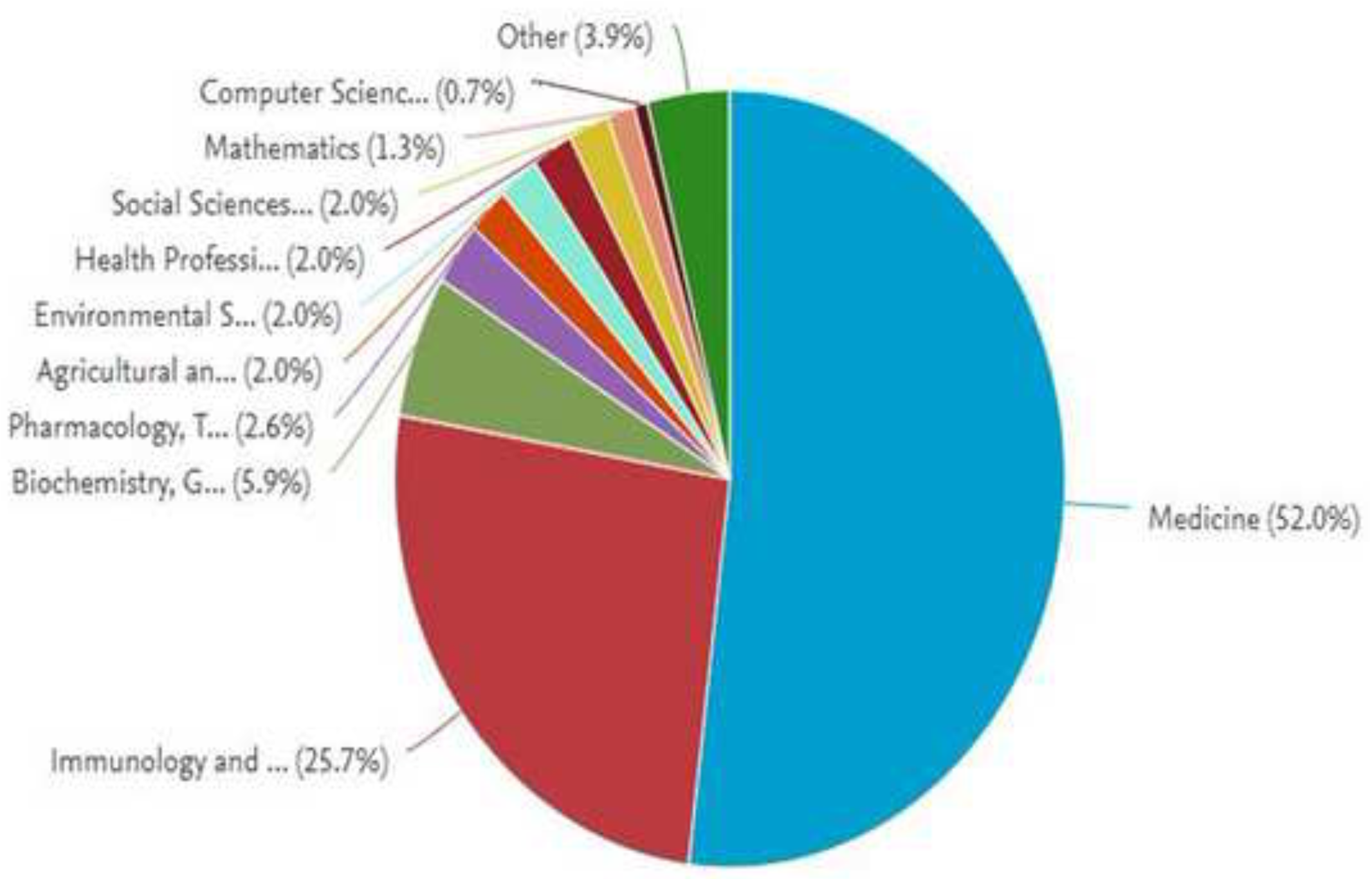
the number of publications of the top 10 popular journals on COVID-19 research.

**Figure 5:**
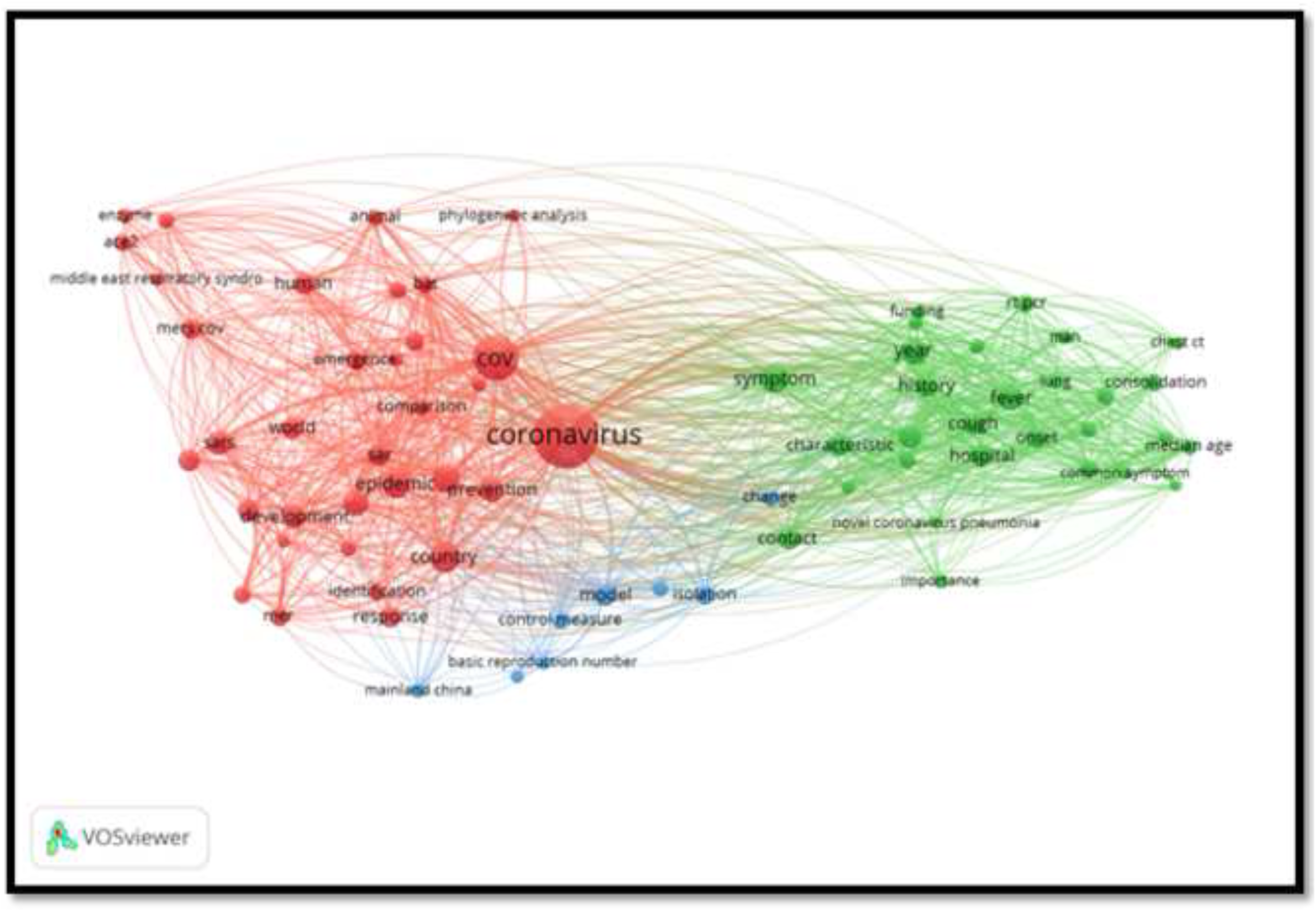
Distribution of document by subject area

### Characteristics of top 10 Covid-19 articles cited most frequently

In total, the top 10 articles contributed 307 citations, accounting for 84.34% of citations related to Covid-19 (**Table 3**). **Hotspots of Research on Covid-19:** VOS viewer was used to analyze keywords extracted from the titles and abstracts of 552 articles included in this study. As a result, 84 keywords, which appeared more than 10 times, were included and shown in the map. These could be stratified into three clusters: High-frequency keywords in cluster 1 were “Coronavirus” (224 times), “Cov” (125 times), and “death” (41 times). For the characteristics-related research in cluster 2, the top keywords are comprised of “patient” (107 times), “symptom” (52 times), and “year”(40 times). In cluster 3 were “Country” 58 times), “epidemic” (49 times), and “sars” (36 times).

**Table 3:** Top 10 Covid-19 research with the most citation frequency.

| No | Title | First author | Journal | IF | Year | Citations | Main Conclusion |
| --- | --- | --- | --- | --- | --- | --- | --- |
| 1 | Clinical features of patients infected with 2019 novel coronavirus in Wuhan, China | Huang, Chaolin | The Lancet | 10.28 | 2020 | 88 | The 2019-nCoV infection caused clusters of severe respiratory illness similar to severe acute respiratory syndrome coronavirus and was associated with ICU admission and high mortality. |
| 2 | A novel coronavirus from patients with pneumonia in China, 2019 | Zhu, Na | New England Journal of Medicine | 70.67 | 2020 | 56 | Different from both MERS-CoV and SARS-CoV, 2019-nCoV is the seventh member of the family of coronaviruses that infect humans. |
| 3 | A familial cluster of pneumonia associated with the 2019 novel coronavirus indicating person-to-person transmission: a study of a family cluster | Chan, Jasper Fw | The Lancet | 10.28 | 2020 | 49 | Person-to-person transmission of this novel coronavirus in hospital and family settings, and the reports of infected travelers in other geographical regions |
| 4 | Epidemiological and clinical characteristics of 99 cases of 2019 novel coronavirus pneumonia in Wuhan, China: a descriptive study | Chen, Nanshan | The Lancet | 10.28 | 2020 | 34 | The 2019-nCoV infection was of clustering onset, is more likely to affect older males with comorbidities, and can result in severe and even fatal respiratory diseases such as acute respiratory distress syndrome. In general, characteristics of patients who died were in line with the MuLBSTA score, an early warning model for predicting mortality in viral pneumonia |
| 5 | Genomic characterisation and epidemiology of 2019 novel coronavirus: implications for virus origins and receptor binding | Lu, Roujian | The Lancet | 10.28 | 2020 | 23 | 2019-nCoV is sufficiently divergent from SARS-CoV to be considered a new human-infecting betacoronavirus. Although their phylogenetic analysis suggests that bats might be the original host of this virus, an animal sold at the seafood market in Wuhan might represent an intermediate host facilitating the emergence of the virus in humans. Importantly, structural analysis suggests that 2019-nCoV might be able to bind to the angiotensin-converting enzyme 2 receptor in humans. |
| 6 | Clinical Characteristics of 138 Hospitalized Patients with 2019 Novel Coronavirus-Infected Pneumonia in Wuhan, China | Wang, Dawei | JAMA - Journal of the American Medical Association | 51.273 | 2020 | 15 | n this single-center case series of 138 hospitalized patients with confirmed NCIP in Wuhan, China, presumed hospital-related transmission of 2019-nCoV was suspected in 41% of patients, 26% of patients received ICU care, and mortality was 4.3%.. |
| 7 | Nowcasting and forecasting the potential domestic and international spread of the 2019-nCoV outbreak originating in Wuhan, China: a modelling study | Wu, J.T | The Lancet | 10.28 | 2020 | 14 | Given that 2019-nCoV is no longer contained within Wuhan, other major Chinese cities are probably sustaining localised outbreaks. Large<br>cities overseas with close transport links to China could also become outbreak epicentres, unless substantial public health interventions at both the population and personal levels are implemented immediately. Independent self-sustaining outbreaks in major cities globally could become inevitable because of substantial exportation of presymptomatic cases and in the absence of large-scale public health interventions. Preparedness plans and mitigation interventions should be readied for quick deployment globally. |
| 8 | Cross-species transmission of the newly identified coronavirus 2019-nCoV | Ji, Wei | Journal of Medical Virology | 2.049 | 2020 | 11 | their findings suggest that 2019-nCoV has most similar genetic information with bat coronavirus and most similar codon usage bias with snake. Taken together, these results suggest that homologous recombination may occur and contribute to the 2019-nCoV cross-species transmission. |
| 9 | Preliminary estimation of the basic reproduction number of novel coronavirus (2019-nCoV) in China, from 2019 to 2020: A data-driven analysis in the early phase of the outbreak | Zhao, Shi | International Journal of Infectious Diseases | 3.538 | 2020 | 9 | The mean estimate of R0 for the 2019-nCoV ranges from 2.24 to 3.58, and is significantly larger than 1.their findings indicate the potential of 2019-nCoV to cause outbreaks |
| 10 | Genomic characterization of the 2019 novel human-pathogenic coronavirus isolated from a patient with atypical pneumonia after visiting Wuhan | Chan, Jasper Fw | Emerging microbes & infections | 4.36 | 2020 | 8 | Remarkably, its orf3b encodes a completely novel short protein. Furthermore, its new orf8 likely encodes a secreted protein with an alpha-helix, following with a beta-sheet(s) containing six strands. Learning from the roles of civet in SARS and camel in MERS, hunting for the animal source of 2019-nCoV and its more ancestral virus would be important for understanding the origin and evolution of this novel lineage B betacoronavirus. These findings provide the basis for starting further studies on the pathogenesis, and optimizing the design of diagnostic, antiviral and vaccination strategies for this emerging infection. |

**Figure 6:** the analysis of key words. The mapping on key words of Covid-19.

For analyzing data in VOS viewer, we used ISI, PubMed, and Scopus as database. The keywords were divided into three clusters: In general, the smaller the distance between two terms, the larger the number of co-occurrences of the terms. A large size of a circle represents that the keyword appears more frequently. The line means that the topics connected on the same line are separated from each other by a comma, a semicolon, or a tab.

## 4. Discussion

In this study, we tried to provide an overview of the world situation in the field of publication of COVID-19. The results indicate that at this ISI, researchers from around the world started publishing the article just on month ago and the number of articles in this field is still growing quickly.

Bibliometric and visualized mapping may quantitatively monitor research performance in science and present predictions [7]. This type of studies had impact on other scientific and professional communities. In the case of antimicrobial resistance surveillance, for example, because real-time surveillance data are often unavailable and limited, scholars have used Scientometrics and found that it provides a fast, reliable, and global overview of research [8]. As a result, Bibliometric studies may be a meaningful reference. In this study, we evaluated Covid-19. Studies with respect to the contributing countries, institutions, journals, and funding sponsors.

In the case of global trends of research on Covid-19, the possibility that the increasing trend will go on longer than that expected from the proposed model, because the application of Covid-19 as a fast-spreading and super-contagious virus might arouse more attention. In terms of country analysis, China published 44 articles and was the leading country in terms of scientific productivity so far. Considering the factors of a large population and GDP, we performed an adjustment and found that published 0.003 articles per trillion GDP and 3.15 articles per million population. Moreover, we found that although the United States published 21 articles and was in second place, its total citations and H-index were 19 and 2, respectively. In terms of journals, we observed that the Euro Surveillance Bulletin “Europeen Sur Les Maladies Transmissibles “European Communicable Disease Bulletin published far more Covid-19 research papers, with 10 articles, than other journals. It was indicated that future development within Covid-19 would likely. Considering the field of research focuses on Covid-19, and the details of the top cited articles, we found that the top studies had been cited 307 times, which indicated that these studies might be classic and fundamental for further studies and should be read by those new to the Field. As shown in Bibliometric mapping of keywords, it was observed that the focus on Covid-19 research is gradually shifting to most basic and experimental studies.

The article titled “Clinical features of patients infected with 2019 novel coronavirus in Wuhan, China” has been cited the most, at 88 times in total was published in “The Lancet” in 2020 [1]. Tis article point the 2019-nCoV infection caused clusters of severe respiratory illness similar to severe acute respiratory syndrome coronavirus and was associated with ICU admission and high mortality. As for the prospective application of the VOS viewer map, we suggest that authors could select research topics from the map and demonstrate its importance as frontier hotspot by the map, and funding agents might be suggested to invest in these orientations.

### Strengths and Limitations

This Bibliometric description and mapping provided a birds-eye view of information on Covid-19 related research for readers to comprehend the history of published Covid-19 articles in just a few minutes. In addition, we evaluated the research strength of countries and institutions, which scholars might refer to in order to find cooperative institutions. During our research using the Scopus database, we tried to guarantee comprehension and objectivity. However, we must consider the following limitations.

Firstly, only publications written in English were included this study, which, inevitably, missed some significant studies on Covid-19 published in other languages. Secondly, as the publication on COVID-19, is growing very quickly, even daily, it is very difficult to extend the findings and generalizing the results. Thirdly, there were still differences between real research conditions and the Bibliometric analysis results, since some recently published papers do not have high citation frequency, as reported by Stephan et al., in Nature [9]. Lastly, the data in this study are open to expansion, with new studies being published each day, and the increasing trend of publication number might go on for longer than is expected from the proposed model.

## Conclusions

The subject of this study was the fast growing publication on COVID-19. Most studies are published in journals with very high impact factors (IFs) and other journals are more interested in this type of research. Future studies, the association of these two topics, and related keywords will be explored to provide more comprehensive results. In addition to the bibliometric analysis of the selected databases, this study focuses solely on published sources; it is recommended that future studies of published sources in Cochrane library, EmBase and other databases also be considered. Overall, since the topic of using information technology in medical virology is widely considered in countries around the world, it seems necessary to provide relevant educational programs to empower medical experts to optimally use these tools in their professional work.

## Data Availability

All data presented in the manuscript.

## Notes

### Competing Interest Statement

The authors have declared no competing interest.

### Funding Statement

None

## References

1. Huang, C., et al., Clinical features of patients infected with 2019 novel coronavirus in Wuhan, China. Lancet, 2020. 395(10223): p. 497–506.

2. Worlometers. 2020 [cited 2020 14 March]; Available from: https://www.worldometers.info/coronavirus/.

3. Lai, C.C., et al., Severe acute respiratory syndrome coronavirus 2 (SARS-CoV-2) and coronavirus disease-2019 (COVID-19): The epidemic and the challenges. Int J Antimicrob Agents, 2020: p. 105924.

4. Cascella, M., et al., Features, Evaluation and Treatment Coronavirus (COVID-19), in StatPearls. 2020, StatPearls Publishing StatPearls Publishing LLC.: Treasure Island (FL).

5. Chen, C., R. Dubin, and M.C. Kim, Emerging trends and new developments in regenerative medicine: a scientometric update (2000 - 2014). Expert Opin Biol Ther, 2014. 14(9): p. 1295–317.

6. Wang, Q., et al., A bibliometric analysis of research on the risk of engineering nanomaterials during 1999-2012. Sci Total Environ, 2014. 473-474: p. 483–9.

7. Tijssen, R.J. and J. Winnink, Twenty-first century macro-trends in the institutional fabric of science: bibliometric monitoring and analysis. Scientometrics, 2016. 109(3): p. 2181–2194.

8. Brandt, C., et al., The bigger picture: the history of antibiotics and antimicrobial resistance displayed by scientometric data. Int J Antimicrob Agents, 2014. 44(5): p. 424–30.

9. Stephan, P., R. Veugelers, and J. Wang, Reviewers are blinkered by bibliometrics. Nature, 2017. 544(7651): p. 411–412.

